# Hydrogen Peroxide Vapor sterilization of N95 respirators for reuse

**DOI:** 10.1101/2020.03.24.20041087

**Authors:** Patrick A. Kenney, Benjamin K. Chan, Kaitlyn Kortright, Margaret Cintron, Nancy Havill, Mark Russi, Jaqueline Epright, Lorraine Lee, Thomas J. Balcezak, Richard A. Martinello

## Abstract

Reprocessing of used N95 respirators may ameliorate supply chain constraints during the COVID-19 pandemic and provide a higher filtration crisis alternative. The FDA Medical Countermeasures Initiative previously funded a study of HP vapor decontamination of respirators using a Clarus C system (Bioquell, Horsham, PA) which normally is used to fumigate hospital rooms. The process preserved respirator function, but it is unknown if HP vapor would be virucidal since respirators have porous fabric that may harbor virus.

We evaluated the virucidal activity of HP vapor using a BQ-50 system (Bioquell, Horsham, PA) after inoculating 3M 1870 N95 respirators (3M, St. Paul, MN) with 3 aerosolized bacteriophage that are a reasonable proxy for SARS-CoV-2. Inoculation resulted in contamination of the respirator with 9.87e4 plaque forming units (PFU) of phage phi-6, 4.17e7 PFU of phage T7 and 1.35e7 PFU of phage T1. Respirators were reprocessed with BQ-50 with a long aeration phase to reduce HP vapors. Virucidal activity was measured by a standard plaquing assay prior to and after sterilization. A single HP vapor cycle resulted in complete eradication of phage from masks (limit of detection 10 PFU, lower than the infectious dose of the majority of respiratory viral pathogens). After 5 cycles, the respirators appeared similar to new with no deformity.

Use of a Bioquell machine can be scaled to permit simultaneous sterilization of a large number of used but otherwise intact respirators. HP vapor reprocessing may ease shortages and provide a higher filtration crisis alternative to non-NIOSH masks.

## Correspondence

The COVID-19 pandemic has led to unprecedented utilization of healthcare resources. There are global shortages of personal protective equipment (PPE), including N95 respirators.^1^ The CDC has modified PPE recommendations and provided “crisis alternate strategies” if the respirator supply is exhausted, including use of masks which are not approved by the National Institute for Occupational Safety and Health (NIOSH) and homemade masks as a last resort.^2,3^ Non-NIOSH and improvised fabric masks provide only marginal protection and are inferior to N95 respirators, a particular concern during the COVID-19 pandemic which has a high rate of healthcare worker infection.^4,5^ Reused respirators may become a reservoir for pathogens, presenting a potential risk.^6,7^ Reprocessing of used N95 respirators may ameliorate supply chain constraints and provide a higher filtration crisis alternative, but it is unknown if effective sterilization can be achieved for a virus without impairing respirator function.

Thermal reprocessing methods deform respirators and alter fit.^8^ Ultraviolet germicidal irradiation of respirators reduces virus viability but efficiency is hampered by shadowing.^9^ Hydrogen peroxide (HP) vapor is virucidal on hard surfaces, and has been shown not to impair respirator performance.^10,11^ HP gas plasma systems are used at hospitals but are ineffective for respirators as filter material absorbs HP vapor; the resultant drop in vapor concentration causes the machine to stop.^10^ The FDA Medical Countermeasures Initiative funded a study of HP vapor decontamination of respirators using a Clarus C system (Bioquell, Horsham, PA) which normally is used to fumigate hospital rooms.^12^ The respirator function was excellent, with no impairment of aerosol collection efficiency or air flow resistance after 50 cycles. Although there was complete inactivation of aerosolized *Geobacillus stearothermophius*, it is unknown if HP vapor would be virucidal since respirators have porous fabric that may harbor virus.

We evaluated the virucidal activity of HP vapor using a BQ-50 system (Bioquell, Horsham, PA) after inoculating 3M 1870 N95 respirators (3M, St. Paul, MN) with 3 aerosolized bacteriophages: T1, T7, and *Pseudomonas* phage phi-6. These phage are a reasonable proxy for SARS-CoV-2 and are an ideal system to study that potentially encompass the spectrum of viral diversity due to notable stability (T1), ease of use (T7) and possession of a viral envelope (phi-6). Aerosols containing phage were generated using a fine mist spray bottle aimed directly at an inverted N95 respirator placed in a 1L flask. Inoculation resulted in contamination of the respirator with 9.87e4 plaque forming units (PFU) of phage phi-6, 4.17e7 PFU of phage T7 and 1.35e7 PFU of phage T1 *per* respirator (performed in triplicate). Concentrations were selected to approximate viral titers necessary for 50% tissue-culture infectious dose (TCID50) of SARS-CoV-2.^13^

Respirators were suspended by their elastic on racks in a 33 m^3^ room and sterilized with BQ-50 using a 10 minute conditioning phase, 30-40 minute gassing phase (varies with humidity and room size) at 16 g/min, 25 minute dwell phase, and a 150 minute aeration phase (varies with number of respirators and room size), with this long duration intended to reduce HP vapors.^10,12^ Half of the respirators had subsequent steam sterilization at 275° F for 5 minutes.

Virucidal activity was measured by a standard plaquing assay on a lawn of host bacteria prior to and after sterilization. Measurements are reported as means across three replicates. A single HP vapor cycle resulted in complete eradication of phage from masks (limit of detection 10 PFU, lower than the infectious dose of the majority of respiratory viral pathogens).^14^ Figure 1. Steam sterilization degraded the respirators and resulted in no additional virucidal activity. After 5 HP vapor cycles, the respirators appeared similar to new with no deformity.

**Figure.**
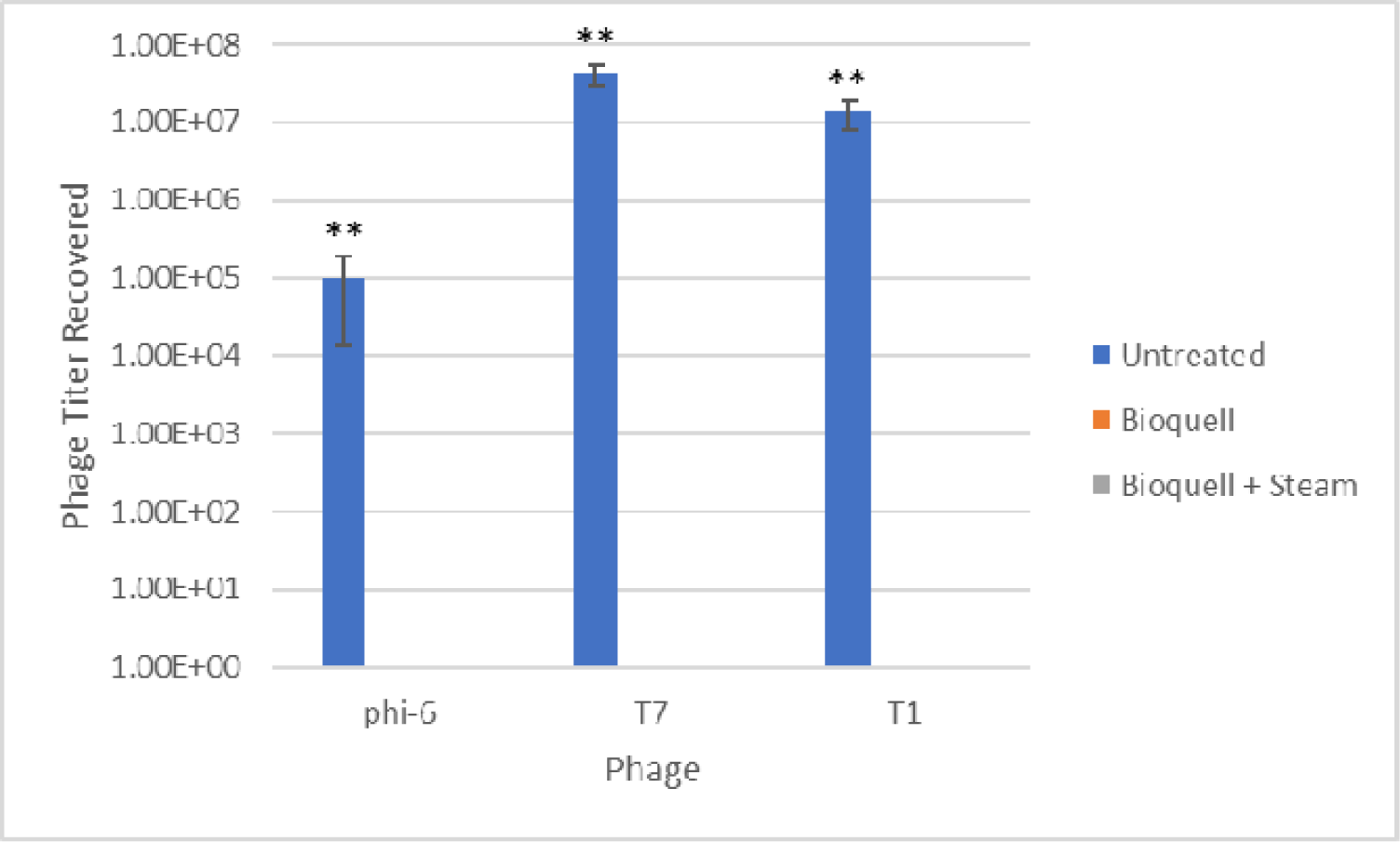
Phage titer recovered from untreated (blue), Bioquell reprocessed (orange), and Bioquell plus steam sterilized (grey) masks. Bioquell reprocessing resulted in no detectable residual phage.

We found that Bioquell HP vapor has high virucidal activity for N95 respirators inoculated with aerosolized virus. Use of a Bioquell machine can be scaled to permit simultaneous sterilization of a large number of used but otherwise intact respirators. HP vapor reprocessing may ease shortages and provide a higher filtration crisis alternative to non-NIOSH masks.

## Data Availability

Data available upon request.

